# The Use of Wastewater Surveillance to Estimate SARS-CoV-2 Fecal Viral Shedding Pattern and Identify Time Periods with Intensified Transmission

**DOI:** 10.1101/2024.08.02.24311410

**Authors:** Wan Yang, Enoma Omoregie, Aaron Olsen, Elizabeth A. Watts, Hilary Parton, Ellen Lee

## Abstract

**Background:** Wastewater-based surveillance is an important tool for monitoring the COVID-19 pandemic. However, it remains challenging to translate wastewater SARS-CoV-2 viral load to infection number, due to unclear shedding patterns in wastewater and potential differences between variants.

**Objectives:** We utilized comprehensive wastewater surveillance data and estimates of infection prevalence (i.e., the source of the viral shedding) available for New York City (NYC) to characterize SARS-CoV-2 fecal shedding pattern over multiple COVID-19 waves.

**Methods:** We collected SARS-CoV-2 viral wastewater measurements in NYC during August 31, 2020 – August 29, 2023 (*N* = 3794 samples). Combining with estimates of infection prevalence (number of infectious individuals including those not detected as cases), we estimated the time-lag, duration, and per-infection fecal shedding rate for the ancestral/Iota, Delta, and Omicron variants, separately. We also developed a procedure to identify occasions with intensified transmission.

**Results:** Models suggested fecal viral shedding likely starts around the same time as and lasts slightly longer than respiratory tract shedding. Estimated fecal viral shedding rate was highest during the ancestral/Iota variant wave, at 1.44 (95% CI: 1.35 – 1.53) billion RNA copies in wastewater per day per infection (measured by RT-qPCR), and decreased by ∼20% and 50-60% during the Delta wave and Omicron period, respectively. We identified around 200 occasions during which the wastewater SARS-CoV-2 viral load exceeded the expected level in any of 14 sewersheds. These anomalies disproportionally occurred during late January, late April - early May, early August, and from late-November to late-December, with frequencies exceeding the expectation assuming random occurrence (*P* < 0.05; bootstrapping test).

**Discussion:** These estimates may be useful in understanding changes in underlying infection rate and help quantify changes in COVID-19 transmission and severity over time. We have also demonstrated that wastewater surveillance data can support the identification of time periods with potentially intensified transmission.

## INTRODUCTION

Since the early phase of the COVID-19 pandemic, studies have reported that wastewater SARS-CoV-2 viral loads often closely track or lead case and/or hospitalization trajectories and, as such, can serve as a cost-effective surveillance tool for monitoring the COVID-19 pandemic.^1–5^ Thus, wastewater-based surveillance systems have been built worldwide on local and national scales. With decreasing clinical testing and genomic sequencing,^6,7^ there has been increased interest in wastewater surveillance, given results are generated independently of clinical testing practice.

Though there are advantages of SARS-CoV-2 wastewater surveillance, a large US national survey of public health agencies completed in 2022 noted the results were often deemed supplementary to surveillance involving clinical laboratory tests.^8^ One of the hurdles is that while the trends could indicate changes in SARS-CoV-2 community circulation, it remains challenging to directly translate wastewater SARS-CoV-2 viral loads to a specific number of infections in the population, due to the unclear fecal viral shedding rate (after accounting for the recovery rate of virus genomes) in wastewater samples. In addition, with the fast emergence and turnover of new SARS-CoV-2 variants, it is unclear how fecal shedding of the virus may have altered over time by variant. To address these questions, we utilize comprehensive wastewater surveillance data and estimates of infection prevalence (i.e., the source of the viral shedding) available for New York City (NYC) to characterize SARS-CoV-2 fecal shedding over multiple COVID-19 pandemic and epidemic waves.

NYC experienced the earliest pandemic wave in the United States (US), and shortly after the initial wave, established a wastewater surveillance program that covers all of its 14 sewersheds which serve over 8 million residents.^2^ Since August 31, 2020, the program has continuously measured SARS-CoV-2 viral load weekly. Independently, we have developed and used a comprehensive model-inference system – calibrated to case, emergency department (ED) visit, and mortality data – to reconstruct the underlying transmission dynamics and estimate key epidemiological characteristics.^9,10^ In particular, the model-inference system estimates the number of infectious individuals including those not detected as cases (i.e., infection prevalence) in each of the city’s 42 neighborhoods during each week since March 1, 2020.^9,10^ Combining the wastewater SARS-CoV-2 viral load data and infection prevalence estimates over a 3-year period (i.e., August 31, 2020 – August 29, 2023), we are able to characterize the viral shedding pattern (i.e., time-lag, duration, and *per-infection* shedding rate) for the ancestral/Iota, Delta, and Omicron variants, separately. We are also able to identify time periods with greater transmission.

## METHODS

### SARS-CoV-2 wastewater surveillance data

The SARS-CoV-2 wastewater surveillance program in NYC started on August 31, 2020. Wastewater samples were taken at each of the city’s 14 wastewater treatment plants, usually twice per week on Sundays and Tuesdays (*N* = 3794 samples; see variations and details in Table S1). SARS-CoV-2 RNA concentration was measured using quantitative reverse transcription polymerase chain reaction (RT-qPCR) assays during August 31, 2020, through April 11, 2023, and reverse transcription digital PCR (RT-dPCR) assays from November 1, 2022, through August 29, 2023. All measurements adjusted for sewershed-specific flow rate and service population size. Specifically, per-capita SARS-CoV-2 viral load (RNA copies per day per population) was computed as the viral concentration measure multiplied by the daily sewage flow rate and then divided by the service population.

For weeks after April 11, 2023, when the samples were measured using RT-dPCR alone, we converted the RT-dPCR measurements to RT-qPCR equivalents, to allow characterization of SARS-CoV-2 viral shedding during the entire Omicron period. Specifically, we first computed the conversion ratio using measurements from November 1, 2022, through April 11, 2023, when both assays were conducted, simply as the mean of all RT-qPCR measurements dividing the mean of all RT-dPCR measurements, during these weeks. We then multiplied the RT-dPCR measurements by the conversion ratio to obtain the converted RT-qPCR equivalents. As an alternative, we stratified the data by sewershed and performed the conversion using sewershed-specific conversion ratios (see Sensitivity Analysis). In addition, the RT-qPCR and RT-dPCR measures differed substantially (by a factor of 16.7 based on the aforementioned overlapping measurements), likely due to difference in methodology.^11^ To facilitate comparison with studies primarily using RT-dPCR, we also converted all RT-qPCR measurements to RT-dPCR equivalents when reporting the viral shedding rates.

### SARS-CoV-2 infection prevalence estimates

Estimated SARS-CoV-2 infection prevalence came from a model-inference system,^12^ independent of the wastewater surveillance data. Briefly, the model-inference system fit a neighborhood-level Susceptible-Exposed-Infectious-(re)Susceptible-Vaccination (SEIRSV) model to age-grouped, neighborhood-specific COVID-19 case, ED visit, and mortality data, accounting for concurrent nonpharmaceutical interventions, vaccinations, under-detection of infection, and seasonal changes. We used the SEIRSV model to explicitly simulate the number of infectious individuals – i.e., anyone who can actively transmit SARS-CoV-2 and infect others regardless of symptoms and test-seeking behaviors – present in the population and estimated this infection prevalence during each week using the full model-inference system using COVID- 19 case, ED visit, and mortality data.^12^ That is, similar to the wastewater SARS-CoV-2 viral loads measuring the total population fecal shedding regardless of clinical testing, estimated infection prevalence here included all individuals actively transmitting SARS-CoV-2 (primarily via shedding from the respiratory tracts) regardless of whether they were detected as cases.

The infection prevalence estimates are United Hospital Fund neighborhood-^13^ and age group specific, and available for each week starting March 1, 2020 (the pandemic onset in NYC) to the week starting August 27, 2023. To match with the sewershed-level wastewater SARS-CoV-2 viral load data, we first mapped each neighborhood (42 in total vs. 14 sewersheds) to the corresponding sewershed based on geolocation; if a neighborhood overlapped multiple sewersheds, we assigned it to the one with the maximal overlap. For each sewershed and week, we then aggregated all estimated infectious individuals from all related neighborhoods.

### Estimating the fecal viral shedding time-lag, duration, and rate

To analyze the fecal viral shedding pattern by variant, we defined three time periods based on data availability and the predominant circulating variant^14^ (i.e., to be more variant-specific): i) the 2^nd^ wave (predominantly the ancestral and Iota variants), from August 31, 2020 (i.e., the first day of wastewater surveillance) through June 26, 2021; ii) the Delta wave (predominantly the Delta variant), from June 27, 2021 (i.e., the first week the share of Delta exceeding 50% among the sequenced specimens) through December 4, 2021; and iii) the Omicron period (predominantly Omicron subvariants and included multiple Omicron-subvariant waves), from December 5, 2021 (i.e., the first week the share of Omicron BA.1 exceeding 25% among the sequenced samples; note that we used a lower threshold here given the milder severity of Omicron BA.1^15^ and thus likely fewer infections detected and sequenced) though August 29, 2023 (i.e., the last wastewater sample during the study period).

SARS-CoV-2 viral load in wastewater represents the pooled fecal shedding of the virus by the population, whereas the infection prevalence represents the proportion of population actively infectious at a given time (i.e., the source of the viral shedding after a potential time-lag). Thus, to estimate the viral shedding rate for each variant (per the time period defined above), we used a linear regression model, accounting for circulating variants and spatial variations by sewershed, per Eq. 1:

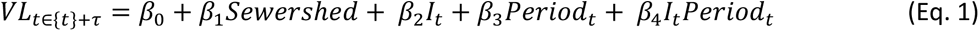

where, *VL_t_*_∈{*t*}+τ_is the wastewater SARS-CoV-2 viral load measured during time-window {*t*}, adjusted by a time-lag or lead of *τ* days (see details below); *Sewershed* is a categorical variable (*Sewershed* = one of the 14 sewersheds in the city) to account for spatial variation; *I_t_* is the infection prevalence estimated for week-*t*; and *Period_t_* represents three epidemic time periods as defined above, included as a proxy for circulating variants during week-*t* (*Period* = 2^nd^ wave, Delta wave, or Omicron period, as defined above). The interaction term *I_t_Period_t_* is included to account for potential nonadditive interaction of the two variables (here, in essence, to allow different viral shedding rates by variant). Per Eq. 1, we computed the estimates of fecal viral shedding rate for each variant using the coefficients β_2_and β_4_.

Given the different surveillance schedules and likely difference between fecal and respiratory viral shedding, we tested three sliding time-windows (i.e., {*t*} in Eq. 1) for matching the wastewater measurements (twice per week, representing fecal shedding) with the infection prevalence estimates (weekly estimates, representing respiratory shedding); specifically, we averaged 2, 3, or 4 consecutive wastewater samples, corresponding to roughly a 1-, 1.5-, or 2- week window, respectively, depending on the wastewater sampling schedule and time-adjustment used. For each time-window {*t*}, to identify a proper time-adjustment (*τ* in Eq. 1), we tested five settings to capture the time difference from becoming infectious via respiratory shedding to fecal shedding per the population-level surveillance data:

1. a 6- to 7-day lead, i.e., the wastewater samples included in time-window {*t*} started from the 1^st^ sample taken *the week before* the infection prevalence estimate; note the 1^st^ sample was taken on Sunday (corresponding to a maximum of 7-day lead) or Monday (corresponding to a maximum of 6-day lead);
2. a 4- to 5-day lead, i.e., the wastewater samples included in time-window {*t*} started from the 2^nd^ sample taken *the week before* the infection prevalence estimate; note the 2^nd^ sample was taken on Tuesday (corresponding to a maximum of 5-day lead) or Wednesday (corresponding to a maximum of 4-day lead);
3. concurrent (no time-difference, *τ*=0), i.e., the wastewater samples included in time- window {*t*} started from the 1^st^ sample taken *the week of* the infection prevalence estimate;
4. a 2- to 3-day lag, i.e., the wastewater samples included in time-window {*t*} started from the 2^nd^ sample taken *the week of* the infection prevalence estimate (a Tuesday sample corresponded to a 2-day lag and a Wednesday sample corresponded to a 3-day lag); and
5. a 7- to 8-day lag, i.e., the wastewater samples included in time-window {*t*} started from the 1^st^ sample taken *the week after* the infection prevalence estimate (a Sunday sample corresponded to a 7-day lag and a Monday sample corresponded to a 8-day lag).

In addition, we performed variant/period-specific analyses for each of the three time-periods defined above, using a similar model form as Eq. 1 but without the terms related to time-period (*Period_t_*). Since the Omicron period included multiple Omicron-subvariant waves, we also performed stratified analyses for the Omicron BA.1 wave (December 5, 2021, through March 4, 2022, i.e., the last week the share of Omicron BA.1 exceeding 50%) and for weeks from March 5, 2022 onwards, separately.

### Identifying timings with higher-than-expected transmission

Visual inspection of the wastewater data showed there were occasional spikes in SARS-CoV-2 viral load, potentially due to intensified transmission. Due to the temporal dynamics and sampling noise, it is challenging to distinguish such potential instances (i.e., a true signal) based on the wastewater data alone. Thus, here we used the infection prevalence estimates, which had accounted for the main underlying transmission factors, to construct the expected SARS-CoV-2 viral load for comparison. Specifically, we first computed the daily infection prevalence using the weekly estimates with a spline smoothing function, and then used those as inputs in Eq. 1 to compute the expected daily SARS-CoV-2 viral load (median and 90% confidence intervals [CI]). Given the large variance in both the infection prevalence estimates and SARS-CoV-2 viral load data, we deemed a wastewater measurement higher than expected, if it was higher than the 95^th^ percentile (i.e., the upper bound of the 90% CI) of the expected SARS-CoV- 2 viral load.

To examine the timing with higher-than-expected SARS-CoV-2 viral load, we grouped the identified anomaly dates into 10-day bins based on calendar time, i.e., the 1^st^ (early), 2^nd^ (mid), and last (late) 10 days of each month; for example, January 1 of 2021, January 5 of 2022, and January 10 of 2023 would all be grouped as “early-January”. This allows recurrent and/or seasonal events to be grouped in the same or nearby bins. To test whether the identified anomalies occurred at random (e.g., due to noise in the data), we further performed a bootstrap test with 5000 random samples. For each bootstrapping set, we randomly sampled *n_anomaly_* (i.e., the number of identified anomalies) dates from the wastewater measurements (*N* = 3794), and then grouped the dates into the same 10-day bins as done for the identified anomalies. We then pooled the 5000 sets together to construct the distribution of each timing. For example, for early-January (the first 10-day calendar bin), with *n_1_*, *n_2_*, …, and *n_5000_* of the dates falling in that bin for the 5000 sets, the likelihood of having *k* (*k*= 0, …, *n_anomaly_*, i.e., from none to all) anomalies during early-January would be:

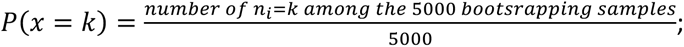

and the likelihood of having *k* or more anomalies during early-January would be:

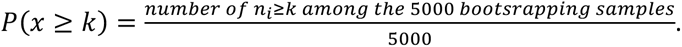

### Sensitivity Analyses

In a first sensitivity analysis, we only included SARS-CoV-2 viral load measured by RT-qPCR (i.e., August 31, 2020– April 11, 2023), to examine if the viral shedding rate estimates were affected by converting RT-dPCR measurements to RT-qPCR equivalents due to changes in testing assays. In a second sensitivity analysis, we included all SARS-CoV-2 viral load measurements but used the sewershed-specific conversion ratios instead of the citywide conversion ratio for all sewersheds.

## RESULTS

### General trends in measured wastewater SARS-CoV-2 viral load and estimated infection prevalence

During the 3-year study period (August 31, 2020 – August 29, 2023), trends in wastewater SARS-CoV-2 viral load were generally consistent with the trends in estimated infection prevalence (Fig 1). Across the 14 NYC sewersheds (Fig 1A), wastewater SARS-CoV-2 viral load tended to rise and fall around the same time (Fig 1B-D and Figs S1-3), indicating epidemic waves were highly synchronized across the city. However, the magnitudes of wastewater SARS-CoV-2 viral load and infection prevalence estimates both varied substantially over time and across sewersheds and may not scale consistently. For example, even though certain sewersheds tended to detect higher SARS-CoV-2 viral loads than others, the rankings changed across different waves (see Fig S1-3, ranked by average viral load). Similar spatial heterogeneity was apparent in the estimated infection prevalence and the discrepancies between wastewater SARS-CoV-2 viral load and estimated infection prevalence appeared larger during the 2^nd^ wave (Fig S1). Such spatial heterogeneity is not unexpected, since several factors such as RNA degradation^16^ and dilution,^16^ and the contribution of infected animals^17^ could all vary by sewershed, and ultimately affect wastewater measurements. In addition, uncertainty in the infection prevalence estimate could also vary by sewershed (e.g., larger uncertainty for those with smaller population size; see, e.g., the wider uncertainty bounds for Oakwood Beach sewershed in Fig S1).

**Fig 1.**
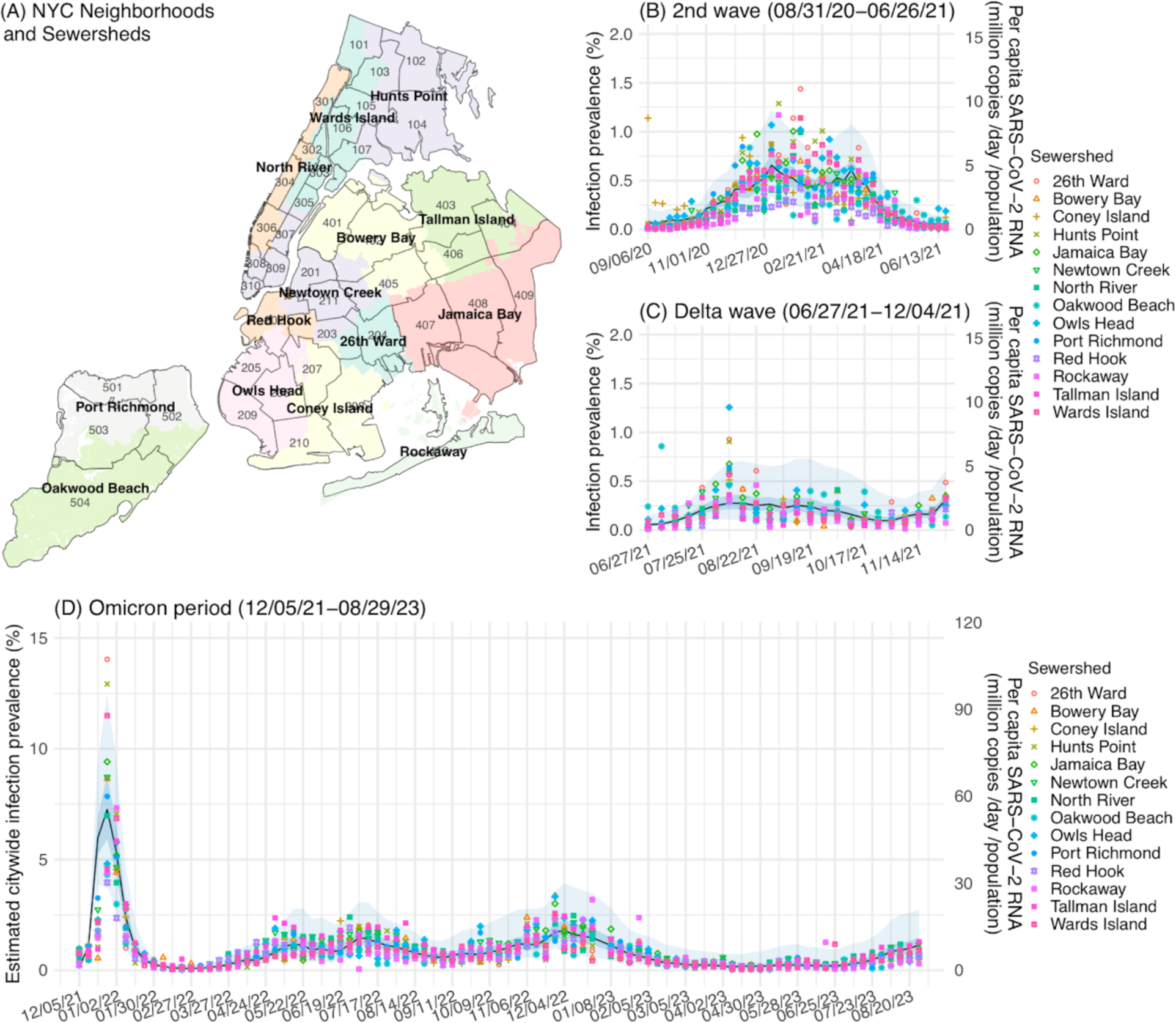
Trends in wastewater SARS-CoV-2 viral load in the 14 sewersheds in NYC. The map in (A) shows 14 sewersheds (delineated by color) and 42 United Hospital Fund neighborhoods (delineated by lines). Dots show the per-capita SARS-CoV-2 viral load in each of the 14 sewersheds (right y-axis, in million copies per day per population by RT-qPCR; color coded per the legend) during the 2^nd^ wave (B), Delta wave (C), and Omicron period (D). For comparison, we overlay the citywide estimates of infection prevalence (left y-axis; blue line = median; darker blue area = 50% CI and lighter blue area = 95% CI).

### Estimated fecal viral shedding patterns

Using the wastewater SARS-CoV-2 viral load data and infection prevalence estimates (i.e., source of fecal viral shedding), we examined fecal viral shedding patterns over the entire study period or stratified by variant/time-period, separately. The estimates are generally consistent (Table 1). Among the 15 combinations of fecal viral shedding time-differences and durations tested, the main model (including all waves) identified concurrent infection prevalence estimates (i.e., no time-difference between becoming infectious via respiratory shedding and fecal shedding) and SARS-CoV-2 viral load aggregated over 3 wastewater samples (2 during the same week and 1 in the beginning of the following week, i.e., a 8- to 9- day-time-interval) as the best setting (highest adjusted R-squared; Fig 2A). Using a 4-5-day-lead and aggregation over 4 wastewater samples (i.e., one sample 4-5 days before, two during, and one 1-2 days after the infection prevalence estimate) led to the second-best model fit (Fig 2A, 2^nd^ dark bar), and was the best setting for the Delta wave and weeks after the BA.1 wave in the stratified analysis (Table 1). Model fit degraded quickly with changing time-differences (both leads and lags), when only 2 (roughly a 1-week duration) or 3 (roughly a 1.5-week duration) wastewater samples were included.

**Fig 2.**
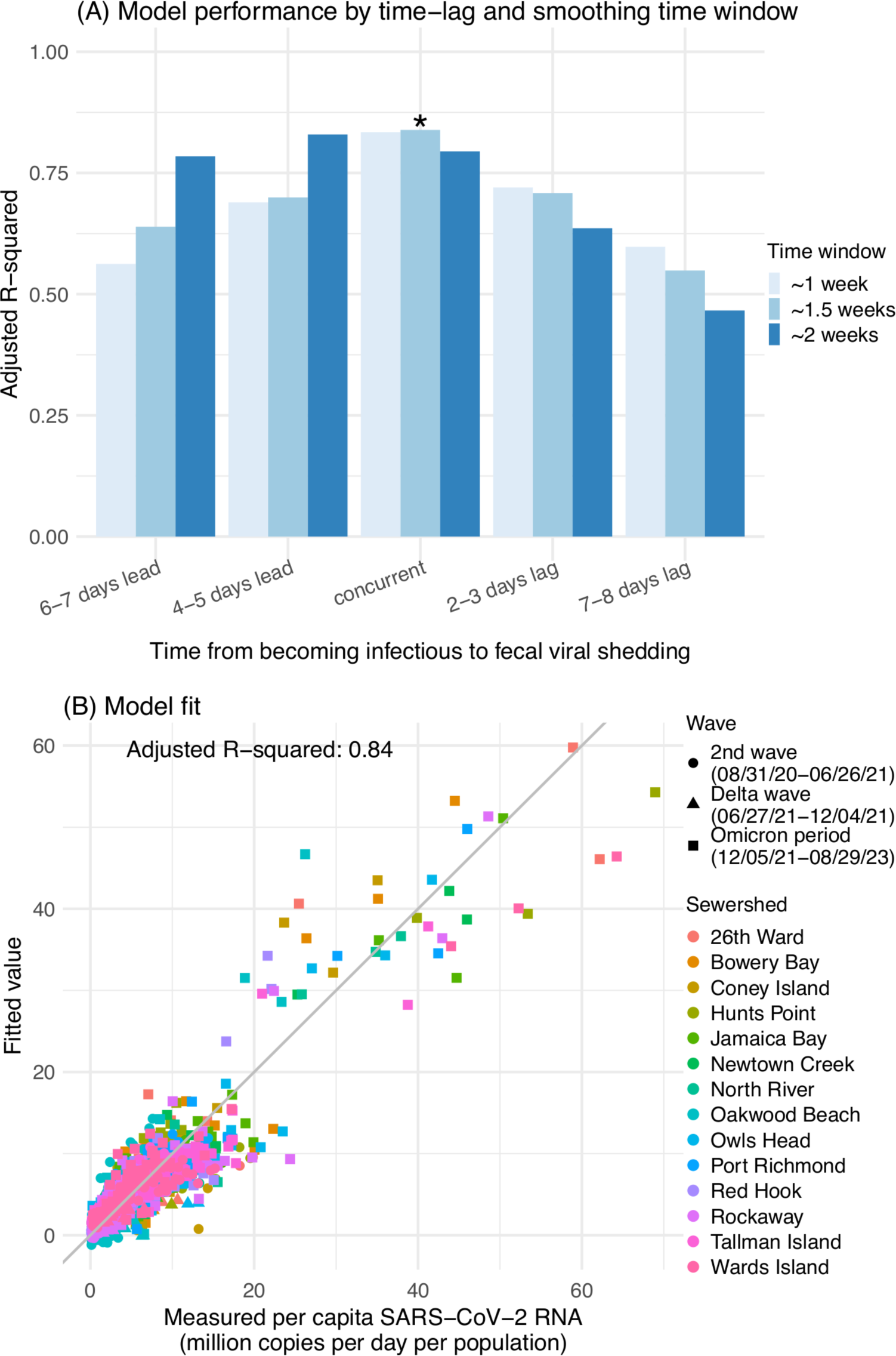
Model fit. (A) shows model performance based on the adjusted R-squared (higher number represents better performance) for different settings of time from becoming infectious to fecal viral shedding and time window of the wastewater samples are aggregated. The asterisk indicates the setting with the highest adjusted R-squared (i.e., best-fit model). (B) shows the model fit compared to the data.

**Table 1.**
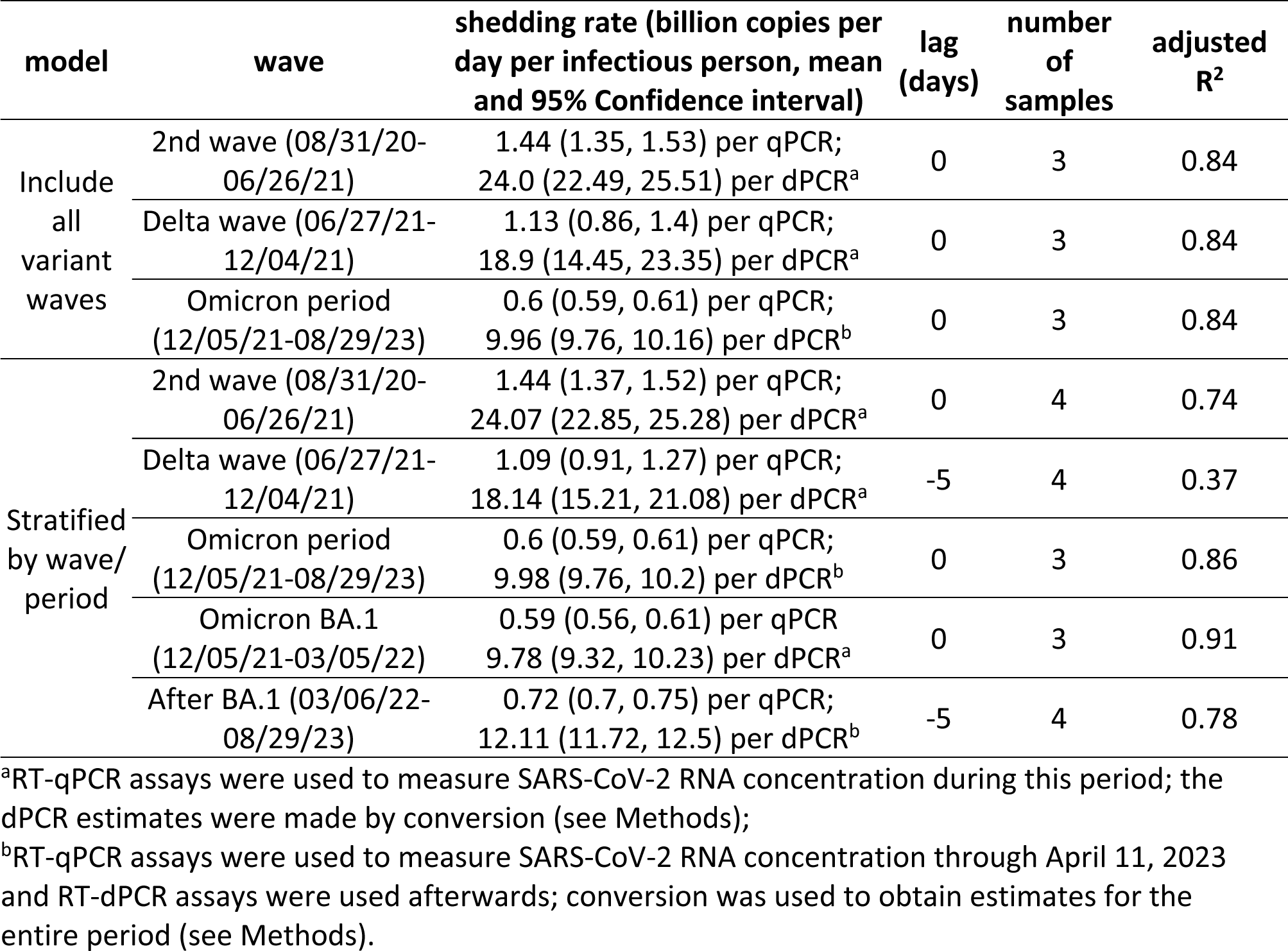
Estimated patterns of SARS-CoV-2 fecal viral shedding in wastewater. Note in this study, SARS-CoV-2 RNA concentration was measured using quantitative reverse transcription polymerase chain reaction (RT-qPCR) assays during August 31, 2020, through April 11, 2023, and reverse transcription digital PCR (RT-dPCR) assays from November 1, 2022, through August 29, 2023. Based on samples tested using both assays, the RT-qPCR and RT-dPCR measures differed by a factor of 16.7. We used this conversion factor to convert measures from the two methods and provide estimates for RT-qPCR and RT-dPCR assays, separately.

Estimated fecal viral shedding rate was highest for infections during the 2^nd^ wave (mostly due to the ancestral and Iota variants), at 1.44 (95% CI: 1.35 – 1.53) billion RNA copies by RT-qPCR in wastewater per day per infectious person [or 24 (95% CI: 22.49 - 25.51) billion RNA copies per RT-dPCR conversion; see Methods]. The estimated rate decreased by ∼20% during the subsequent Delta wave and by 50-60% during the Omicron period (Table 1). Importantly, we note the lower estimates for Delta and Omicron may in part reflect reduced shedding among vaccinees and recoverees, in addition to variant-specific variations.

### Timings with higher-than-expected transmission

The infection prevalence estimates have accounted for the general transmission factors (here, population-level mobility, vaccinations, variant-specific properties, and seasonal risk of infection; see Methods), but may have not fully accounted for activities such as increased gatherings during certain time-periods that might increase transmission. In contrast, wastewater SARS-CoV-2 viral load is a composite measure of all transmission events. Thus, comparison of these two quantities could support identification of such events. Following a procedure designed per this mechanism (see Methods), we identified 198 occasions where wastewater SARS-CoV-2 viral loads exceeded the expected levels in any of the 14 sewersheds (see Fig 3A for identified anomalies for Newtown Creek, the sewershed with the largest service population). These anomalies disproportionally occurred during late January, late April - early May, early August, and mid-November to late-December (Fig 3B), with frequencies exceeding the expectation assuming random occurrence. Among the 5000 bootstrapping sets, none had as many or more anomalies as observed in early August or late November (*P* = 0) and less than 5% had as many or more anomalies as observed in late January, late April, early May, late November, and late December (*P* < 0.05 for all these calendar bins; Table S2).

**Fig 3.**
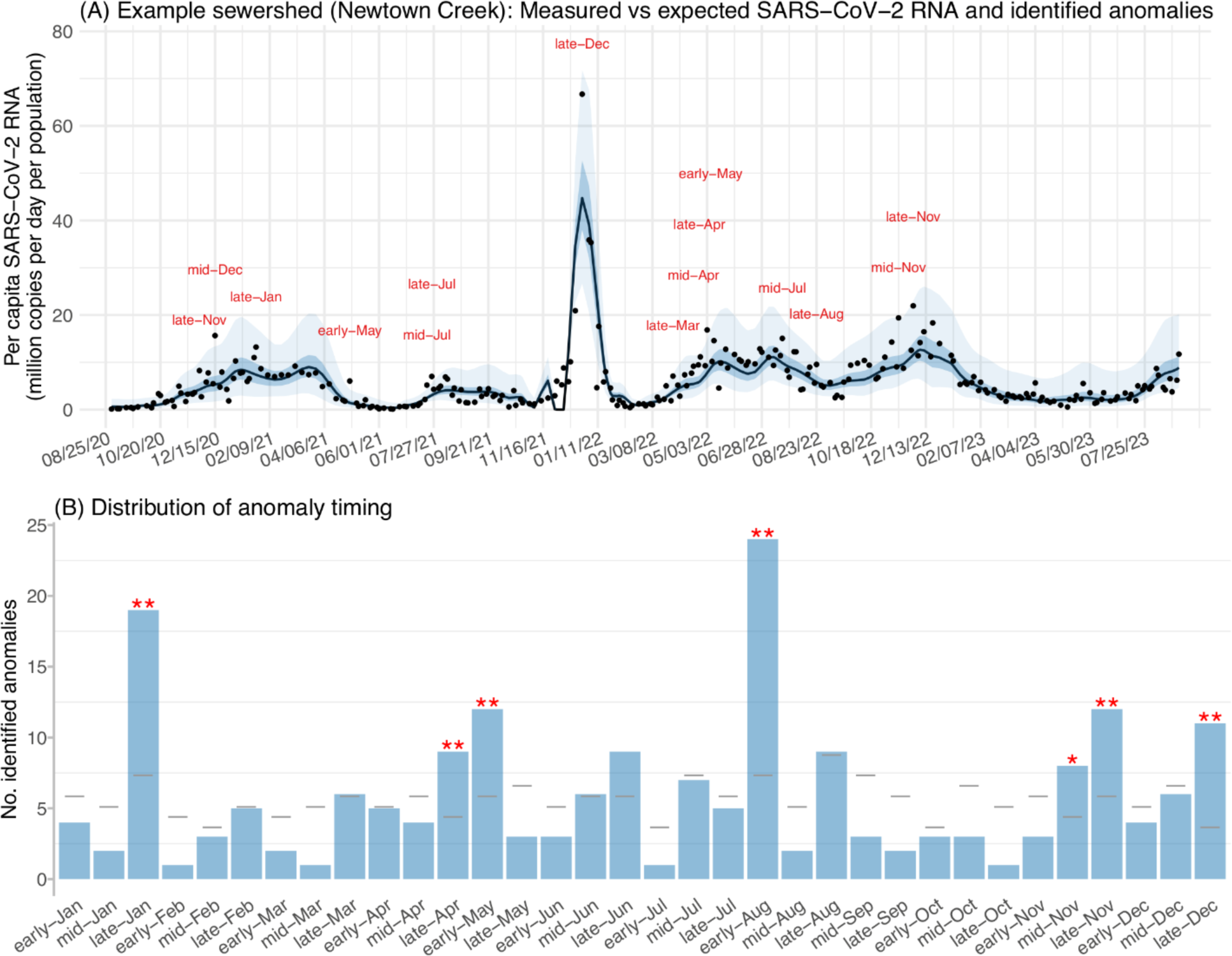
Identified time-periods with intensified transmission in any of 14 NYC sewersheds. (A) shows an example of the measured (dots) and expected wastewater SARS-CoV-2 viral load (blue line = median; darker blue area = 50% CI and lighter blue area = 95% CI), and identified anomalies with SARS-CoV-2 viral load exceeding the expected (red labels). (B) shows the distribution of all identified anomalies. Asterisks indicate time-periods that exceeded the expected wastewater SARS-CoV-2 viral load with a frequency higher than chance assuming random occurrence per a bootstrapping test (* for *P* < 0.1 and ** for *P* < 0.05). Spatial distribution of the anomalies is shown in Fig S4.

### Sensitivity analyses

Results from the two sensitivity analyses are consistent with the main analysis. In the 1^st^ sensitivity analysis (i.e., using SARS-CoV-2 viral load measured by RT-qPCR alone, for a shorter study period from 8/31/20 to 4/11/23), similar fecal viral shedding rates were estimated (Table S3). The 2^nd^ sensitivity analysis (using sewershed-specific conversion ratios to convert the RT-dPCR measurements after 4/11/23, same study period as the main analysis) estimated the same fecal viral shedding rates as the main analysis, and identified three additional anomalies (i.e., 1 in late-January, 1 in mid-August, and 1 in early-July).

## DISCUSSION

Wastewater surveillance can be a valuable tool for monitoring SARS-CoV-2 circulation in the population. To further develop understanding of wastewater surveillance data, we have combined independent model-inference estimates of infection prevalence to characterize fecal viral shedding patterns for multiple major SARS-CoV-2 variants. Using NYC as an example, we have also demonstrated that these data and estimates can support the identification of time periods with potentially intensified transmission.

Importantly, here we examined how wastewater SARS-CoV-2 viral shedding is related to estimated infection prevalence, rather than health outcomes as in previous studies. This choice could lead to certain apparent differences but has several advantages. First, previous studies have reported detection of SARS-CoV-2 in wastewater (e.g., an increase in viral load, or the presence of a new variant) several days ahead of the detection of cases, hospitalizations, or deaths, due to the delay in health outcomes.^18–20^ Here, infection prevalence is a proxy of respiratory tract shedding, which could precede fecal viral shedding. Indeed, we found wastewater SARS-CoV-2 viral loads measured round 1.5 week of the infection prevalence estimate afforded the best model fit (Table 1). This finding suggests that fecal viral shedding likely starts around the same time an individual becomes infectious and lasts slightly longer than the shedding from respiratory tract. Consistent with our finding, studies have shown that fecal SARS-CoV-2 RNA was detectable in patients within the first week of COVID-19 diagnosis and could last longer than respiratory shedding.^16,21^

Second, case-, hospitalization-, or death-to-wastewater-viral-load ratio could decrease with increased vaccinations/reinfections and circulation of milder variants (e.g., Omicron) due to reduced severity or testing, and such reductions have been reported.^19,22^ In contrast, as our estimates included all infections regardless of severity or testing, the infection-to-wastewater- viral-load ratio (roughly, the inverse of estimated per-infection fecal viral shedding rate; Table 1) is relatively stable during each variant wave. For example, the wave-stratified analysis estimated similar fecal viral shedding rates for the BA.1 wave and weeks after BA.1 (Table 1). Importantly, using the infection prevalence estimates, we are able to quantify the fecal viral shedding rate for each major SARS-CoV-2 variant/time-period (Table 1). These estimates can be used to account for changes in underlying infection rate during this study period (e.g. converting wastewater SARS-CoV-2 viral loads to infection prevalence per Table 1) and help examine changes in COVID-19 severity (e.g., changes in hospitalization rate and infection-fatality risk). Such wastewater-viral-load and infection-based estimates may be more accurate than case-based measures, which are subject to test-seeking biases.

Third, previous studies have measured viral loads in clinical samples from the respiratory tract. Based on the reported cycle threshold (CT) values, the respiratory tract viral load was higher in Delta and Omicron infections than the ancestral variant,^23–28^ consistent with the higher infectiousness of these variants of concern. In contrast, fecal viral shedding is not a main mode of transmission,^29,30^ and here using variant circulation time-period as a proxy, we estimate that the fecal viral shedding rate was the highest for the ancestral/Iota variants, followed by Delta (∼20% lower), and then Omicron (∼50-60% lower; Table 1). Early studies of ancestral SARS-CoV- 2 infections found that patients with diarrhea shed more viruses than patients without diarrhea (see, e.g., a review in ref. ^16^), suggesting fecal viral shedding may be associated with diarrhea. In addition, studies found that vaccinations reduced the number of diarrhea episodes,^31^ and that rates of diarrhea were highest among patients infected with the ancestral SARS-CoV-2, followed by patients infected with Delta and then Omicron.^32,33^ Our estimates are consistent with the fecal viral shedding studies,^16,31–33^ and support a difference in viral load between SARS-CoV-2 fecal shedding and respiratory tract shedding, in addition to the timing difference noted above.

In addition to characterizing SARS-CoV-2 fecal viral shedding pattern, we are also able to identify certain time-periods with intensified transmission. In NYC, analysis based on calendar timing showed likely intensified transmission during late-November through December (Fig 3B). Increased transmission also occurred during early August and late January. It is possible that other factors such as travel, holidays, or specific COVID-19 sub-variants could help explain these periods of intensified transmission, but further investigation is needed to determine their impact.

Lastly, we note several limitations. First, given the biweekly sampling dates for wastewater and weekly estimates for infection prevalence, we were unable to test finer time-differences and durations when examining SARS-CoV-2 fecal shedding pattern. Second, the estimates here were based on population data and thus represent an average of all individuals undergoing different disease stages in the population. As such, the estimated fecal shedding duration may be shorter than that reported in studies based on individual patient data (e.g., days or weeks after respiratory tract samples became negative^16^). Third, our infection prevalence estimates have accounted for the main transmission factors, through the information encapsulated in the COVID-19 case, ED visit, and mortality data used for model estimation. Thus, the expected SARS-CoV-2 viral load constructed using these estimates and in turn the identified anomalies are both conservative estimates and may have missed additional anomalies. In addition, wastewater collected from sewersheds may represent individuals who are residents of NYC as well as outside NYC, while infection prevalence estimates are based on NYC residents only.

In summary, we have characterized the fecal viral shedding pattern of SARS-CoV-2 in wastewater in New York City from 2020-2023. These estimates can be used to account for changes in underlying infection rate and help more accurately quantify changes in COVID-19 transmission and severity over time. We have also demonstrated that wastewater surveillance data combined with model-inference estimates can support the identification of time-periods that potentially intensify transmission. Additional studies are needed to better understand these periods and the potential to mitigate SARS-Cov-2 transmission.

## Data Availability

The SARS-CoV-2/COVID-19 cases, emergency department visits, mortality, and wastewater surveillance data were used with permission under a Data Use and Nondisclosure Agreement between the NYC DOHMH and Columbia University.

## Acknowledgements

This study was supported by the National Institute of Allergy and Infectious Diseases (AI175747) and Centers for Disease Control and Prevention (CDC) and the Council of State and Territorial Epidemiologists (CSTE; contract no.: NU38OT00297). The authors thank Lauren Firestein for overseeing the data use agreement and facilitating data sharing for this project; Ramona Lall for providing syndromic surveillance emergency department data; Wenhui Li for providing COVID- 19-associated mortality data; Iris Cheng for providing immunization data; Jubayer Ahmed, Nelson De La Cruz, and Brandon Nguyen for managing and providing wastewater data; the NYC DOHMH Respiratory Pathogens data team for overarching data management and provision of data for this project; and Shama Ahuja, Sharon Greene, Scott Harper, Elizabeth Luoma, Ulrike Siemetzki-Kapoor, Celia Quinn, and Faten Taki for their input on this manuscript.

## Author contributions

WY designed the study, performed the analysis, and wrote the first draft; EO, AO, and EAW oversaw provision of the SARS-CoV-2 wastewater surveillance data; HP and EL oversaw provision of the COVID-19 case and emergency department visit data. All authors contributed to the final draft.

## Supplement Tables and Figures

**Fig S1.**
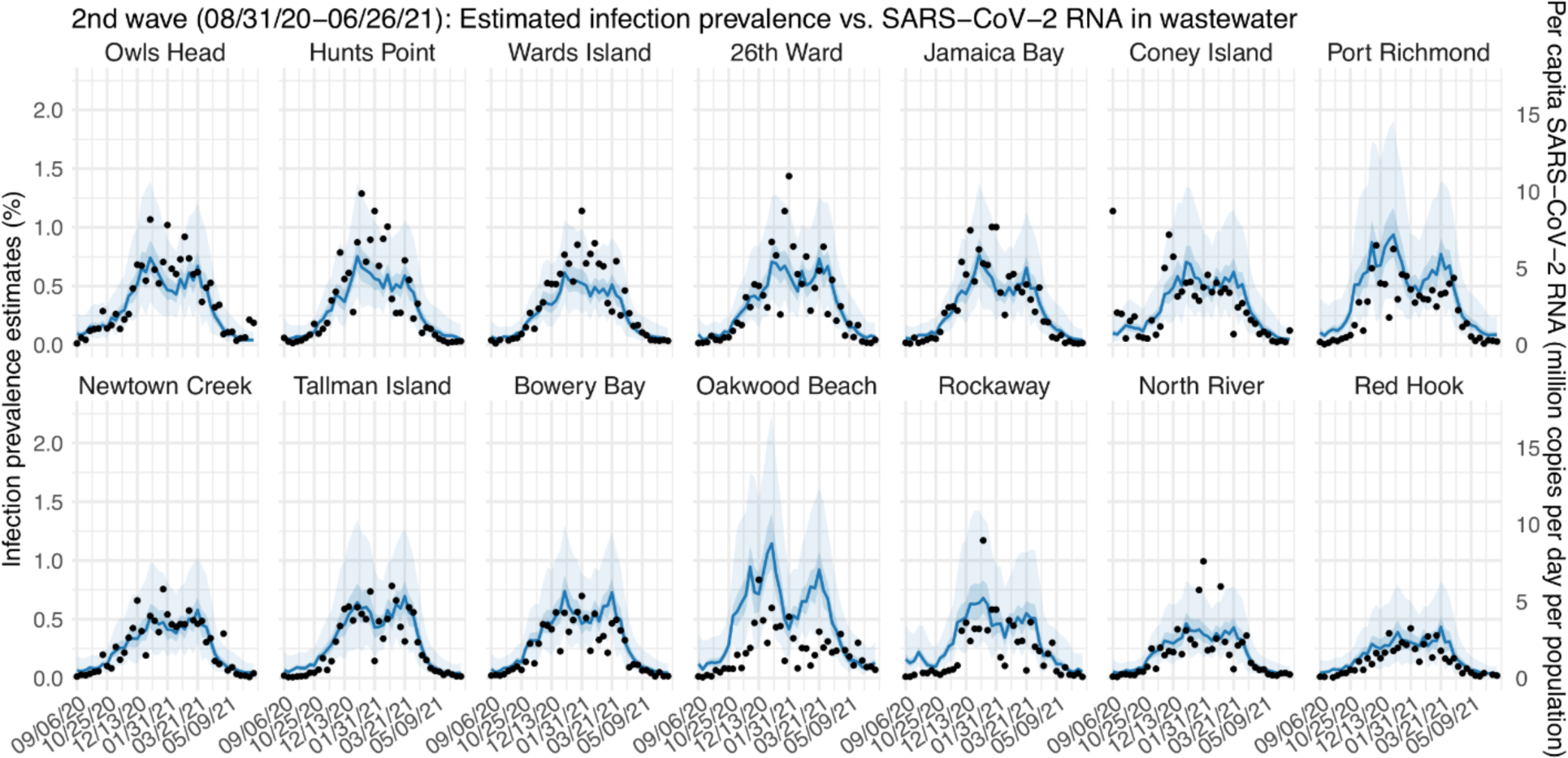
Per-capita wastewater SARS-CoV-2 viral load in each of the 14 NYC sewersheds during the 2^nd^ wave. Dots showed aggregated wastewater SARS-CoV-2 viral load for each week. For comparison, we overlay the corresponding estimates of infection prevalence (blue line = median; darker blue area = 50% CI and lighter blue area = 95% CI). The sewersheds are ordered by the mean viral load during this time period (from the highest to the lowest).

**Fig S2.**
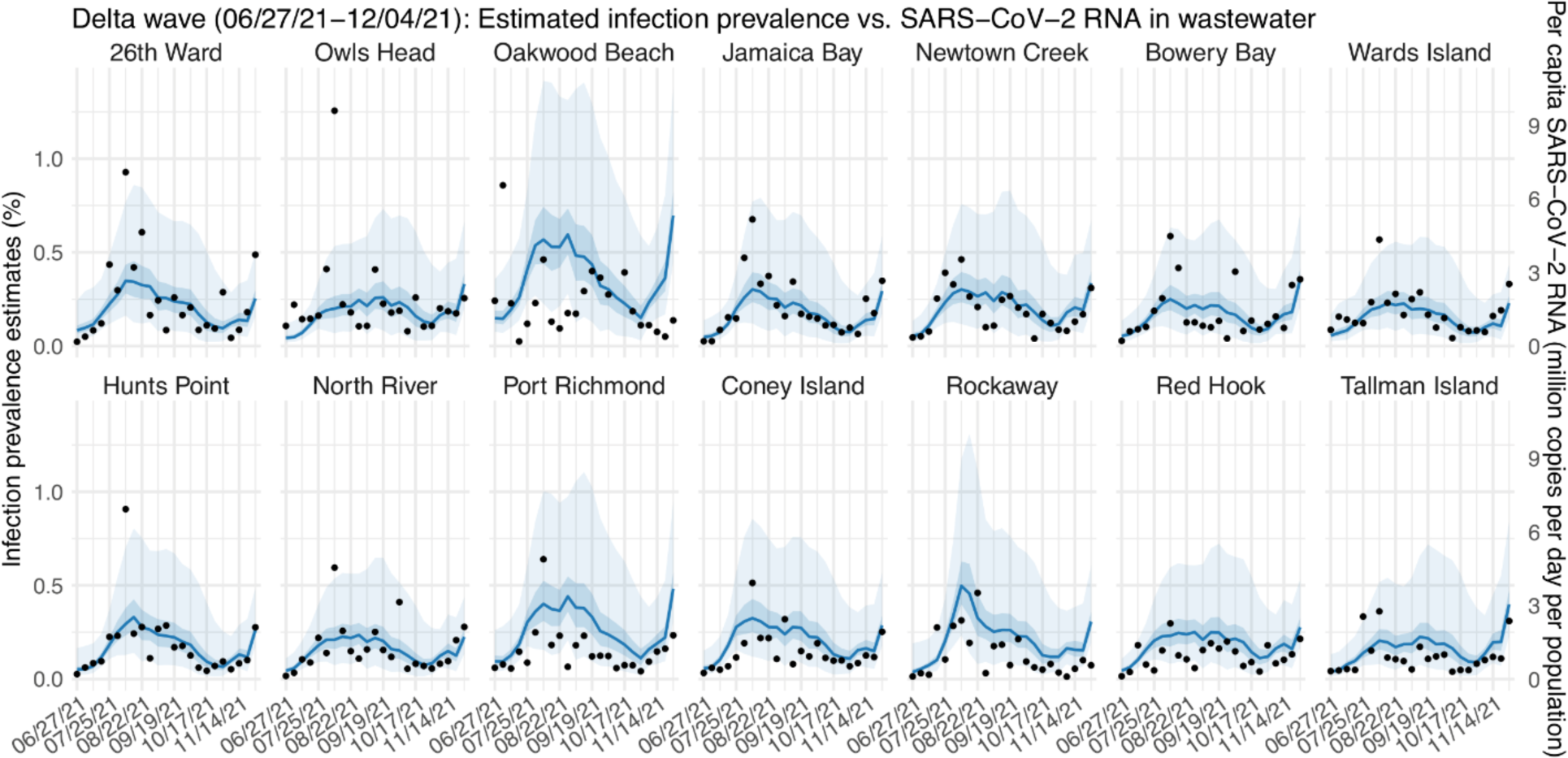
Per-capita wastewater SARS-CoV-2 viral load in each of the 14 NYC sewersheds during the Delta wave. Dots showed aggregated wastewater SARS-CoV-2 viral load for each week. For comparison, we overlay the corresponding estimates of infection prevalence (blue line = median; darker blue area = 50% CI and lighter blue area = 95% CI). The sewersheds are ordered by the mean viral load during this time period (from the highest to the lowest).

**Fig S3.**
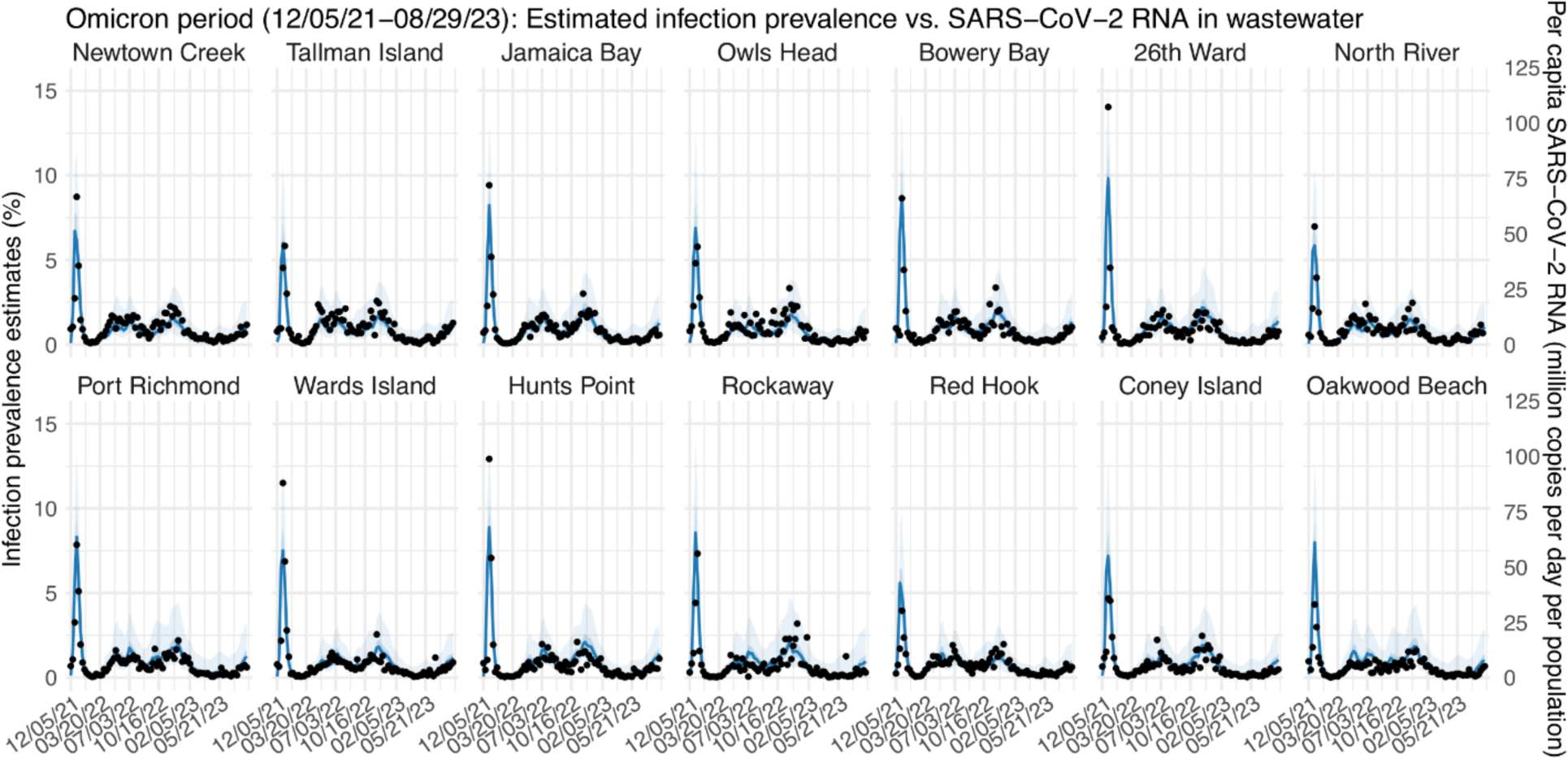
Per-capita wastewater SARS-CoV-2 viral load in each of the 14 NYC sewersheds during the Omicron period. Dots showed aggregated wastewater SARS-CoV-2 viral load for each week. For comparison, we overlay the corresponding estimates of infection prevalence (blue line = median; darker blue area = 50% CI and lighter blue area = 95% CI). The sewersheds are ordered by the mean viral load during this time period (from the highest to the lowest).

**Fig S4.**
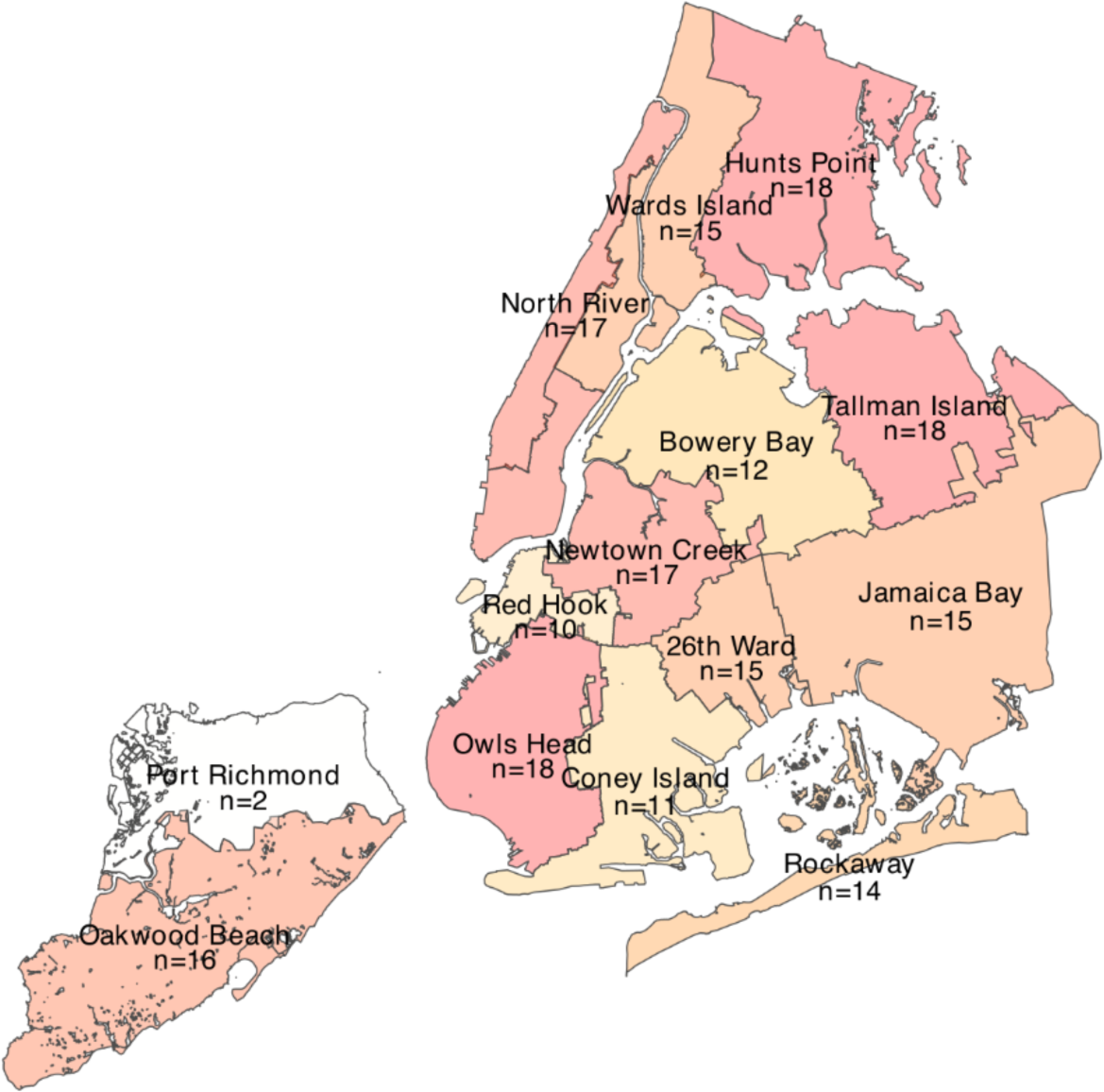
The total number of anomalies identified for each sewershed during the study period (n; see numbers in the map; darker colors indicate larger numbers).

**Table S1.** Summary statistics for the wastewater samples.

| statistics | value | n | percentage |
| --- | --- | --- | --- |
| Total No. of samples | - | 3794 | 100% |
| Day of sampling | Sunday | 1834 | 48.3% |
| Day of sampling | Tuesday | 1736 | 45.8% |
| Day of sampling | Wednesday | 126 | 3.3% |
| Day of sampling | Monday | 98 | 2.6% |
| Sampling frequency | 2 per week | 1610 | 73.7% |
| Sampling frequency | 1 per week | 574 | 26.3% |
| Calendar time | late-Aug | 168 | 4.4% |
| Calendar time | early-Aug | 140 | 3.7% |
| Calendar time | late-Jan | 140 | 3.7% |
| Calendar time | mid-Jul | 140 | 3.7% |
| Calendar time | mid-Sep | 140 | 3.7% |
| Calendar time | late-May | 126 | 3.3% |
| Calendar time | mid-Dec | 126 | 3.3% |
| Calendar time | mid-Oct | 126 | 3.3% |
| Calendar time | early-Jan | 112 | 3% |
| Calendar time | early-May | 112 | 3% |
| Calendar time | early-Nov | 112 | 3% |
| Calendar time | late-Jul | 112 | 3% |
| Calendar time | late-Jun | 112 | 3% |
| Calendar time | late-Mar | 112 | 3% |
| Calendar time | late-Nov | 112 | 3% |
| Calendar time | late-Sep | 112 | 3% |
| Calendar time | mid-Apr | 112 | 3% |
| Calendar time | mid-Jun | 112 | 3% |
| Calendar time | early-Apr | 98 | 2.6% |
| Calendar time | early-Dec | 98 | 2.6% |
| Calendar time | early-Jun | 98 | 2.6% |
| Calendar time | late-Feb | 98 | 2.6% |
| Calendar time | late-Oct | 98 | 2.6% |
| Calendar time | mid-Aug | 98 | 2.6% |
| Calendar time | mid-Jan | 98 | 2.6% |
| Calendar time | mid-Mar | 98 | 2.6% |
| Calendar time | mid-May | 98 | 2.6% |
| Calendar time | early-Feb | 84 | 2.2% |
| Calendar time | early-Mar | 84 | 2.2% |
| Calendar time | late-Apr | 84 | 2.2% |
| Calendar time | mid-Nov | 84 | 2.2% |
| Calendar time | early-Jul | 70 | 1.8% |
| Calendar time | early-Oct | 70 | 1.8% |
| Calendar time | early-Sep | 70 | 1.8% |
| Calendar time | late-Dec | 70 | 1.8% |
| Calendar time | mid-Feb | 70 | 1.8% |

**Table S2.** Likelihood of having the same or higher frequency of anomalies during each calendar time as the observed, based on bootstrapping.

| **timing** | **number anomalies during this time** | **total number of anomalies** | **observed frequency** | **P-value: probability based on bootstrapping** |
| --- | --- | --- | --- | --- |
| early-Aug | 24 | 198 | 0.1212 | 0.0000 |
| late-Jan | 19 | 198 | 0.0960 | 0.0002 |
| late-Dec | 11 | 198 | 0.0556 | 0.0008 |
| late-Nov | 12 | 198 | 0.0606 | 0.0124 |
| early-May | 12 | 198 | 0.0606 | 0.0138 |
| late-Apr | 9 | 198 | 0.0455 | 0.0286 |
| mid-Nov | 8 | 198 | 0.0404 | 0.0680 |
| late-Jun | 9 | 198 | 0.0455 | 0.1292 |
| late-Aug | 9 | 198 | 0.0455 | 0.5122 |
| mid-Jun | 6 | 198 | 0.0303 | 0.5364 |
| late-Mar | 6 | 198 | 0.0303 | 0.5380 |
| late-Feb | 5 | 198 | 0.0253 | 0.5850 |
| early-Apr | 5 | 198 | 0.0253 | 0.5860 |
| mid-Jul | 7 | 198 | 0.0354 | 0.6134 |
| mid-Dec | 6 | 198 | 0.0303 | 0.6546 |
| late-Jul | 5 | 198 | 0.0253 | 0.7060 |
| mid-Feb | 3 | 198 | 0.0152 | 0.7068 |
| early-Oct | 3 | 198 | 0.0152 | 0.7148 |
| early-Dec | 4 | 198 | 0.0202 | 0.7582 |
| early-Jan | 4 | 198 | 0.0202 | 0.8450 |
| mid-Apr | 4 | 198 | 0.0202 | 0.8490 |
| early-Jun | 3 | 198 | 0.0152 | 0.8900 |
| early-Nov | 3 | 198 | 0.0152 | 0.9384 |
| early-Mar | 2 | 198 | 0.0101 | 0.9386 |
| late-May | 3 | 198 | 0.0152 | 0.9634 |
| mid-Oct | 3 | 198 | 0.0152 | 0.9634 |
| mid-Jan | 2 | 198 | 0.0101 | 0.9686 |
| mid-Aug | 2 | 198 | 0.0101 | 0.9720 |
| early-Jul | 1 | 198 | 0.0051 | 0.9772 |
| mid-Sep | 3 | 198 | 0.0152 | 0.9788 |
| late-Sep | 2 | 198 | 0.0101 | 0.9830 |
| early-Feb | 1 | 198 | 0.0051 | 0.9888 |
| late-Oct | 1 | 198 | 0.0051 | 0.9924 |
| mid-Mar | 1 | 198 | 0.0051 | 0.9956 |

**Table S3.**
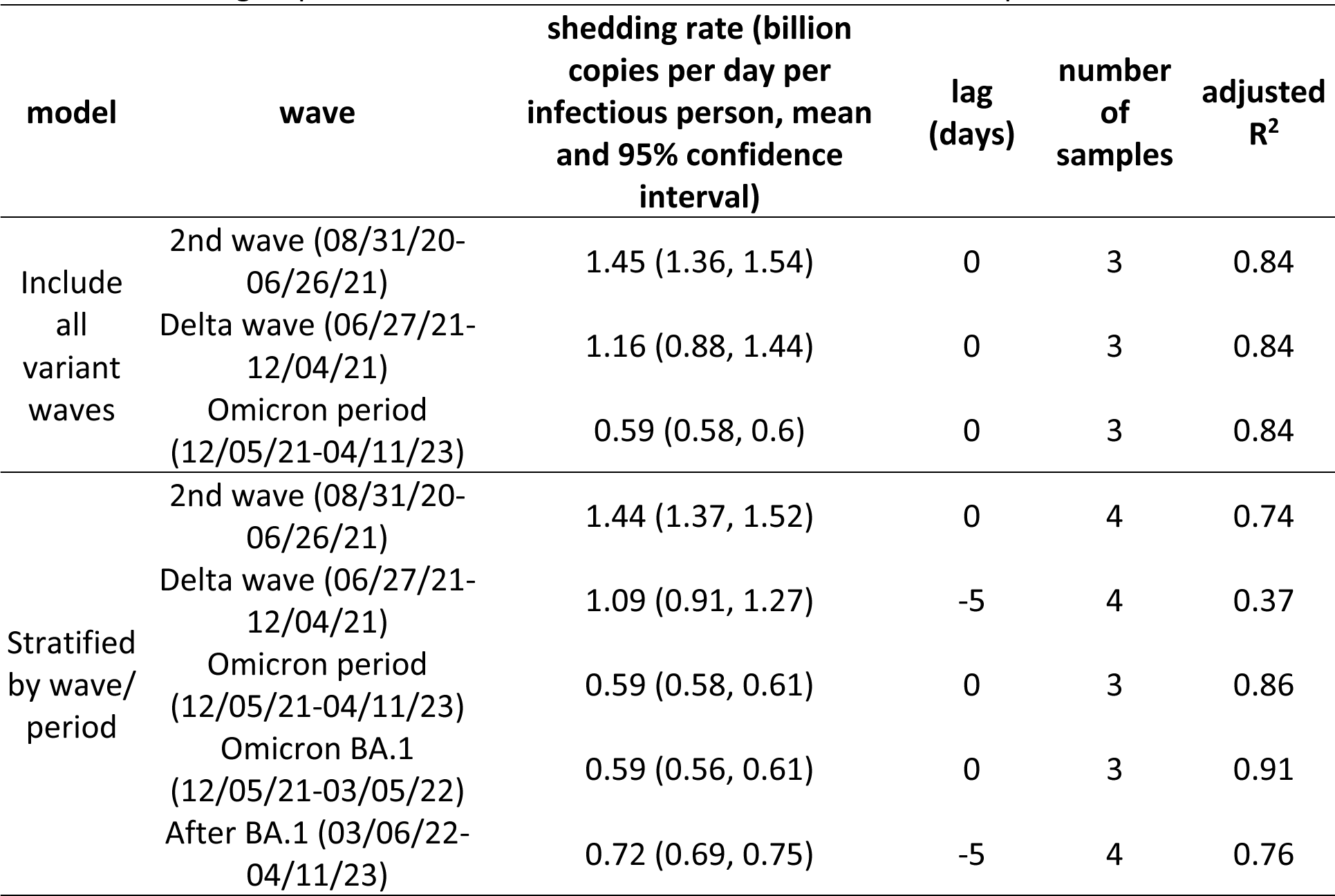
Estimated patterns of SARS-CoV-2 fecal viral shedding in wastewater, using RT-qPCR data alone through April 11, 2023. All estimates here are based on RT-qPCR measures.

| model | wave | shedding rate (billion<br>copies per day per<br>infectious person, mean<br>and 95% confidence<br>interval) | lag<br>(days) | number<br>of<br>samples | adjusted<br>R<sup>2</sup> |
| --- | --- | --- | --- | --- | --- |
| Include<br>all<br>variant<br>waves | 2nd wave (08/31/20-06/26/21) | 1.45 (1.36, 1.54) | 0 | 3 | 0.84 |
|  | Delta wave (06/27/21-12/04/21) | 1.16 (0.88, 1.44) | 0 | 3 | 0.84 |
|  | Omicron period<br>(12/05/21-04/11/23) | 0.59 (0.58, 0.6) | 0 | 3 | 0.84 |
| Stratified<br>by wave/<br>period | 2nd wave (08/31/20-06/26/21) | 1.44 (1.37, 1.52) | 0 | 4 | 0.74 |
|  | Delta wave (06/27/21-12/04/21) | 1.09 (0.91, 1.27) | -5 | 4 | 0.37 |
|  | Omicron period<br>(12/05/21-04/11/23) | 0.59 (0.58, 0.61) | 0 | 3 | 0.86 |
|  | Omicron BA.1<br>(12/05/21-03/05/22) | 0.59 (0.56, 0.61) | 0 | 3 | 0.91 |
|  | After BA.1 (03/06/22-04/11/23) | 0.72 (0.69, 0.75) | -5 | 4 | 0.76 |

